# Analysis of the *All of Us* Research Program Researcher Workbench Workspaces

**DOI:** 10.1101/2022.05.12.22274998

**Authors:** Rhea P. Kerawala, Paula F. King, Kelsey Ross Mayo, Julia Moore Vogel, the All of Us Research Program

**Affiliations:** The Scripps Research Institute, La Jolla, California, United States of America; Vanderbilt Institute for Clinical and Translational Research (VICTR), Vanderbilt University Medical Center, Nashville, Tennessee, United States of America

## Abstract

The *All of Us* Research Program, through the *All of Us* Research Hub platform’s Researcher Workbench, provides researchers and citizen scientists with access to a broad dataset of surveys, electronic health records, physical measurements, genetic data, and Fitbit device wearable data. *All of Us* has a goal of recruiting a minimum of one million participants and aims to capture the diversity of individuals in the United States. The *All of Us* Research Hub platform includes a Research Projects Directory on its website, displaying researcher-provided descriptions of each active workspace on the Researcher Workbench. Inspired by the initial work completed by *All of Us* investigators in 2019, these workspace descriptions were analyzed with a new methodology. Genetic, methods and validation studies, and educational research purposes were associated with disease-focused research, as compared to non-disease focused research. Of all the population categories of interest, only race and ethnicity were associated with disease-focused research. Further categorization of the disease-focused workspaces revealed the top five disease categories: cardiovascular disease, brain and mental health disorders, cancer and benign tumors, diabetes, and immunology-related conditions. Athena OHDSI catalog terms were sorted and helped classify the workspaces by each disease category. Subcategory distribution for the cancer, genetic, and cardiovascular disease-related conditions was examined as well. This framework has the potential to be used for continued longitudinal analysis of the workspaces and continued learnings regarding the importance of disease-focused research in public health. Additionally, as workspace descriptions are created at project initiation, this can provide us with a leading-edge indication of researcher interest in the *All of Us* Research Program data.

## INTRODUCTION

### Background

The *All of Us* Research Program (*AoU*RP) is a collaborative, multi-institution effort to accelerate health research, including by redefining data usage. With a goal of recruiting one million or more participants living in the United States, the National Institutes of Health (NIH) sponsored program includes a comprehensive approach to researching precision medicine, allowing researchers and citizen scientists to explore electronic health record (EHR), survey, physical measurement, genomic, and wearable device data for use in their own research projects. Among the many *AoU*RP program partners is The Participant Center, which consists of the Scripps Research Translational Institute and its partners, and is tasked with directly enrolling participants in the *AoU*RP [1].

Vanderbilt University Medical Center, along with the Broad Institute of Harvard and MIT and Verily Life Sciences, lead the Data and Research Center (DRC). The DRC is responsible for secure housing and curation of participant data and making it available to the research community via the *All of Us* Research Hub [2]. As stated in the *All of Us* Data Access Framework policy, researchers are responsible for declaring their research purpose when they create a workspace to analyze *AoU*RP data [3]. This information is made accessible publicly on the Research Projects directory [4]. Workspace descriptions contain valuable insights which can help inform *AoU*RP staff, participants, funding agencies, and other researchers how the data is currently being used. This information can also be used to evaluate the ability of the *AoU*RP to catalyze research focused on understanding and improving health outcomes for historically underrepresented communities [5]. To our knowledge, this analysis provides the first summary of active *All of Us* research projects from workspace descriptions. Using the workspace descriptions, the research was categorized as diseased-focused or non-diseased-focused to evaluate the workspaces that mentioned various disease categories, including genomic research. In addition, we present descriptive statistics on which diseases were most commonly studied during the first year that data was available for researcher use, in support of the *AoU*RP program goal to understand and facilitate research that will be of importance to participants. Through the researcher workspaces, a more systematic approach to the workspace description data is possible, encompassing the interplay between health habits, lifestyle choices, and disease.

### Goals

The objective for this initial analysis is to examine how information from the workspace descriptions can be used to summarize *AoU*RP research activity for the public. The statistics from the first year of workspace data serve as a framework for characterizing various aspects of the *All of Us* research ecosystem, and underscores the importance of the *AoU*RP in the study of human disease in a diverse population [6]. Subsequent, more focused analysis of workspace data from June 2021 emphasizes evaluating disease-focused (DFR) and non-disease focused (non-DFR) research as a major point of differentiation between the types of workspaces and relevant study topics, such as research purposes and study populations of interest. As the datasets were cumulative, methodology was refined after the initial analysis. Thus, development, refinement, and dissemination of these methods for identifying research activity information from these workspace descriptions was one of our primary goals. In addition, institutional affiliation was investigated to identify where particular workspace descriptions are created geographically, and if there were any patterns associated with affiliation to specific institutions. A focus on genetic research workspaces was also performed to investigate researcher interest into the use of genetic data that the *AoU*RP would eventually provide. In categorizing workspace descriptions by disease conditions, we aimed to identify common patterns or current interests in healthcare research and potentially determine how this data is most useful to the research the *AoU*RP enables.

It was anticipated that analysis of workspace description data can help inform future research projects and present both qualitative and quantitative descriptions of research activity that may be of interest not only to participants, but also to funding agencies, and to the public. In particular, we hypothesize that research purpose and populations of interest data can elucidate the way the *AoU*RP data was being used at the time of this report, which could inform what types of data various researchers are interested in and the potential applications of that data.

## MATERIALS AND METHODS

In this analysis, the DRC provided descriptions for active researcher workspaces from three time points, April 21st, 2021, June 7th, 2021, and June 30th, 2021, as .csv files designated cumulative datasets A, B, and C, respectively. Using the initial analysis of dataset A, we quantified workspace creation and modification times and preferred data types, and created the disease categories manually, as identified in Table 1. These categories were based on a previous *AoU*RP publication to maintain consistency, and adapted using the most common workspace focus areas and major disease areas [6]. Using the Athena OHDSI catalog to determine which terms would be searched for in each category, we confirmed that they were broad enough to encompass several related conditions within each disease category [7]..

**Table 1.** Disease Categories.

|  |
| --- |
| Cardiovascular Disease and Related Disorders |
| Cancer and Benign Tumors |
| COVID-19 |
| Dermatological Disorders |
| Diabetes |
| Gastrointestinal (GI) and Endocrine Disorders |
| Hypertension |
| Immunology |
| Mental Health and Brain-Related Disorders |
| Musculoskeletal Conditions |
| Reproduction |
| Respiratory Ailments |
| Sleep-Related Disorders |
| Obesity |
| Other |

*Manually curated, these categories were determined based on the workspace descriptions of dataset A, and partially based on previous categorization [6]. The other category was used to group the workspaces that did not fit within one of the listed categories.*

Data counts from dataset A, B, and C were compared to determine if there were any differences in the distribution of workspace descriptions over the past few months of data collection (April-June 2021). Statistical analysis was performed using Fisher’s Exact test for categorical data with a p-value cutoff of 0.05 given the relatively small sample size. This was the only statistical test used for consistency, although it was especially important for evaluating genetic research due to the smaller sample size. Distribution of cardiovascular disease and cancer workspaces was also examined, given the significant number of workspace descriptions that sought to investigate these topics, as found in initial analysis.

Using R, filters were applied for the various parameters in sorting workspace data, including deduplication and determining aggregate counts. For each of three datasets, these counts are summarized in Table 2. DFR was determined by filtering workspaces that had “checked” the disease research box at the time that particular workspace was created. This type of filter was applied to research purposes and populations of interest, including subgroups for race and ethnicity as well. Major R packages used included *dplyr* and *tidyverse*. The primary focus of each workspace was found by searching for terms belonging to each disease category, primarily sourced from the Athena OHDSI catalog, particularly ICD and SNOMED condition codes [7]. The number of workspaces attributed to each institution and frequency of that count was also noted. The steps of this workflow are summarized graphically in Figure 1.

**Table 2.**
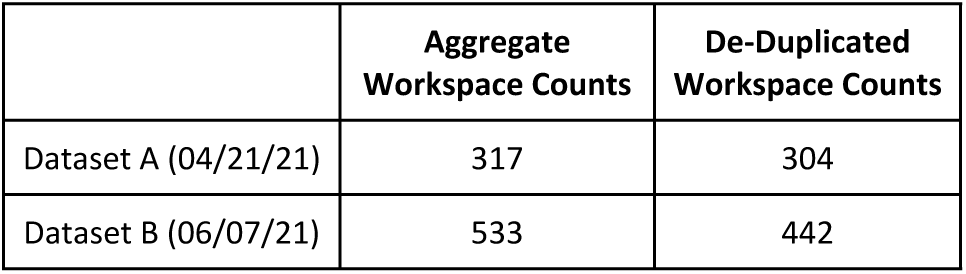
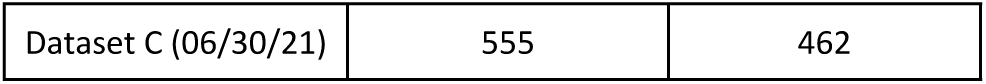
Workspace Counts in each Dataset (pre- and post-de-duplication)

|  | Aggregate<br>Workspace Counts | De-Duplicated<br>Workspace Counts |
| --- | --- | --- |
| Dataset A (04/21/21) | 317 | 304 |
| Dataset B (06/07/21) | 533 | 442 |
| Dataset C (06/30/21) | 555 | 462 |

**Figure 1.**
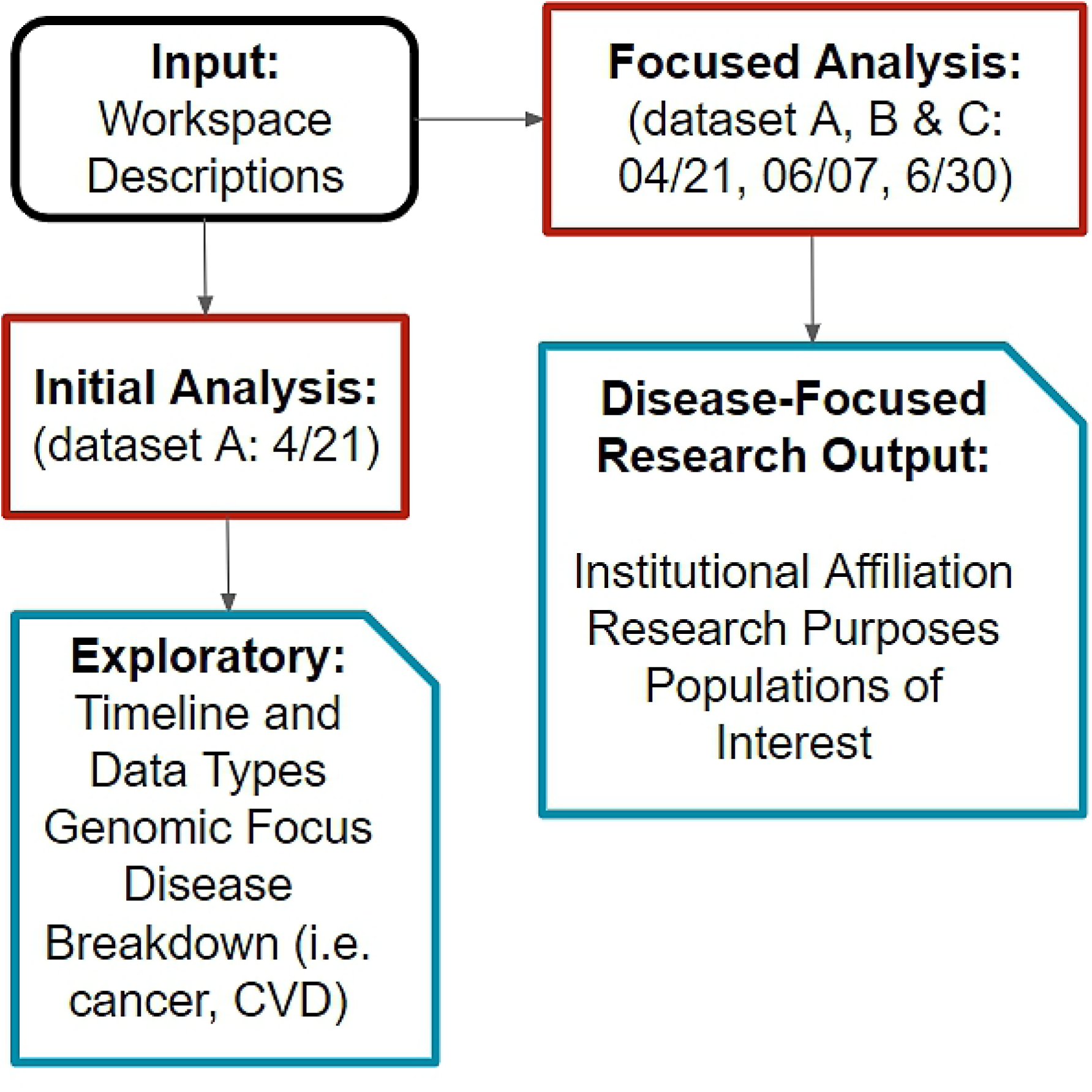
Analysis Workflow. This diagram demonstrates the steps taken to examine the workspace descriptions. While dataset A was initially explored manually, all three datasets were analyzed in R, comparing DFR to non-DFR.

One source of bias that should be acknowledged is that study intent, disease of interest, research purpose, and populations of interest are subject to change over the course of the study. While DRC is notified of researcher updates to the workspace descriptions, this study is limited to the descriptions when the data is pulled for analysis at each time point. For example, with dataset C, any updates made to those workspaces after June 30^th^, was not part of the results, and has to be followed up with future research. As study description and other related information is part of the researcher’s workspace, it is updated at the discretion of the researcher on the Researcher Workbench, and finally in the Research Projects Directory [3].

## RESULTS

Over time, the most common disease categories of the DFR workspaces were cardiovascular-related concerns with around 15.4% of workspaces, mental health and brain-related disorders with about 13.9% of workspaces, cancer and benign tumors at a lower 10.7%, diabetes at 9.29%, immunology at 8.2%,, and reproduction at 6.7% (Figure 2). COVID-19 was studied somewhat significantly, with 3.29% of the workspaces in dataset A and 4.11% of the workspaces in dataset C that indicated an interest in the topic, but was not one of the highest focuses. Interestingly, the distribution of the most commonly studied diseases did not change significantly between the 3-month period evaluated in this study.

**Figure 2.**
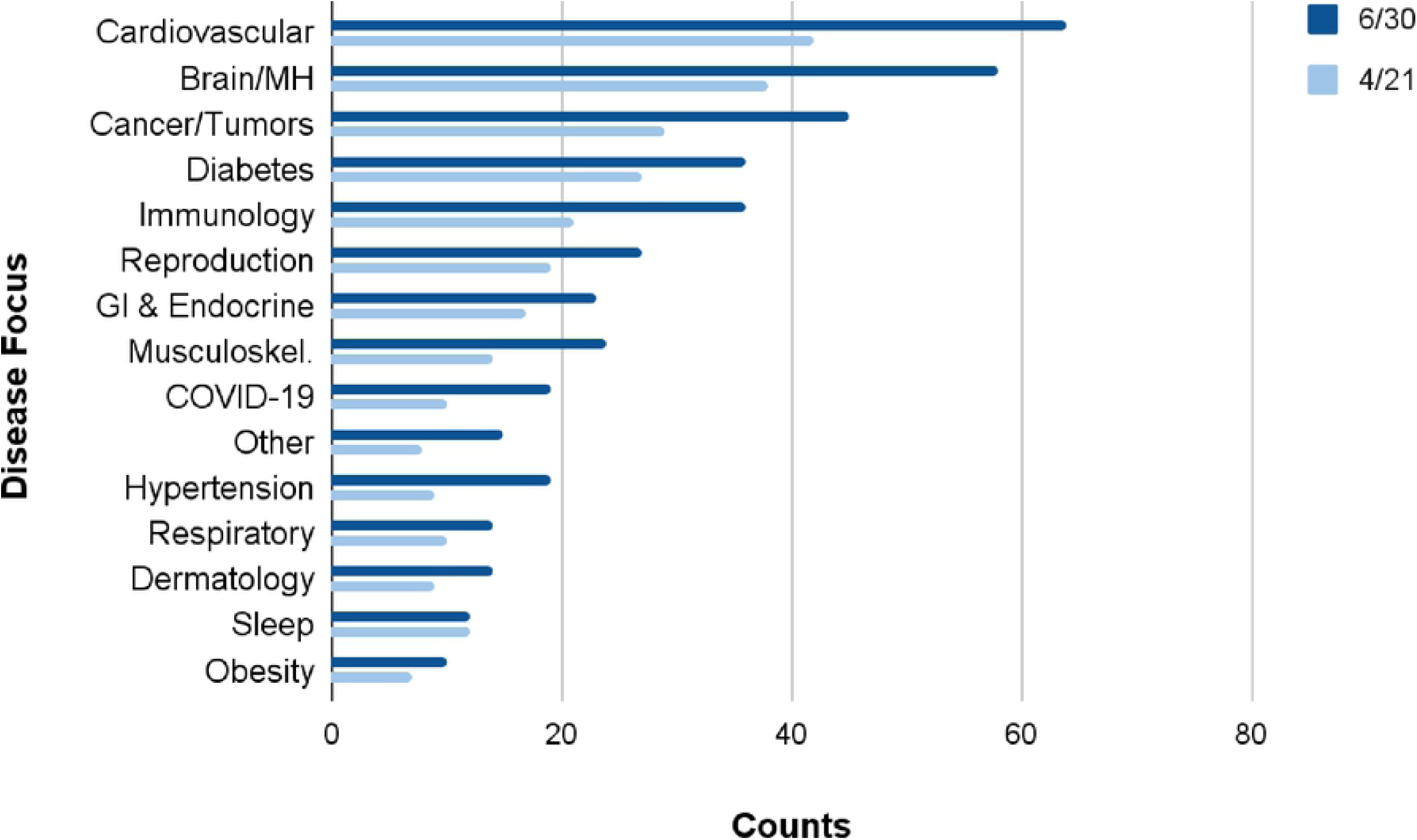
Disease Categories. Over time, the top 5 disease categories were cardiovascular disease and related ailments, followed by brain and mental health disorders, cancer and tumors, diabetes, immunology. The distribution of the categories was consistent, with none being longitudinally significantly different (p > 0.05).

The research purposes that were most common in the most recent data for DFR included population and public health, genetic, and social behavioral research, indicating an expected focus on healthcare applications (Figure 3). Comparatively, non-DFR projects were broader in focus, with the most common being population and public health, education, and methods development and validation. The proportion of workspaces devoted to genetic, methods and validation, education, and other research purposes were statistically significant when comparing disease and non-disease focused research (*p* < 0.05).

**Figure 3.**
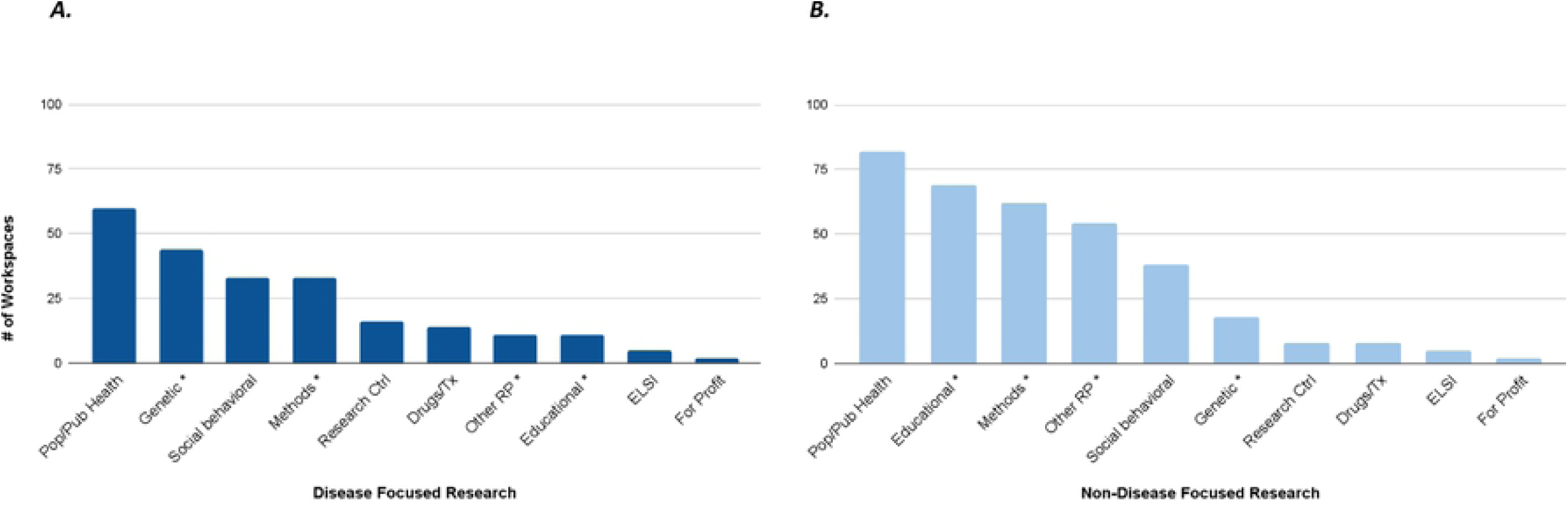
Comparing Research Purposes. Disease-Focused Research (Panel A) compared to non-Disease Focused Research (Panel B). Asterisks are shown for the statistically different research purposes(p < 0.05).

Notably, genetic data from *All of Us* participants were not available to researchers at the time of this analysis, which demonstrates workspace information can be used as an indication of researcher future intent and interest. Despite the unavailability of genetic data to researchers at the time of this analysis, 20.37% of DFR workspaces and 7.32% non-DFR workspaces actually described genetic and/or genomic research within the workspace research project descriptions (Figure 3). This highlights the importance of genomic data to *AoU*RP researchers. In examining disease categories for the genetic workspaces only, the top research focuses again were cardiovascular-related diseases and mental health and brain-related conditions, with the rest following the trend of the overall disease categories. Over time, examining the distribution of the workspace descriptions within each of the disease categories did not result in any statistically significant differences between datasets A and C (*p* < 0.05) (Figure S1).

For the populations who have been historically underrepresented in biomedical research in dataset C, race and ethnicity and age were the two most commonly studied categories, with race and ethnicity being significantly higher in DFR compared to non-DFR on a statistical level. Among the historically underrepresented groups available to study, disability status was the least commonly studied. The most commonly studied race and ethnicity categories were Black, African, and African American; Hispanic, Latino, and Spanish; and Asian American populations (Figure 4). Overall, this evidence supports the *AoU*RP’ s progress towards its goal of focusing research on underrepresented populations [4]. It is important to note the Alaska Native/American Indian participant data was not available for research at the time of this study, but is planned for future release in accordance with recommendations from the *All of Us* Research Program Tribal Consultation [8]. Workspaces that selected this subcategory actually show only an intention to study Alaska Native/American Indian data when it becomes available. There is a significant focus on older adults between the ages of 65 to over 75, which is promising when considering previous challenges with recruitment and retention [9]. There was no statistically significant difference between the DFR and non-DFR categories for age groups.

**Figure 4.**
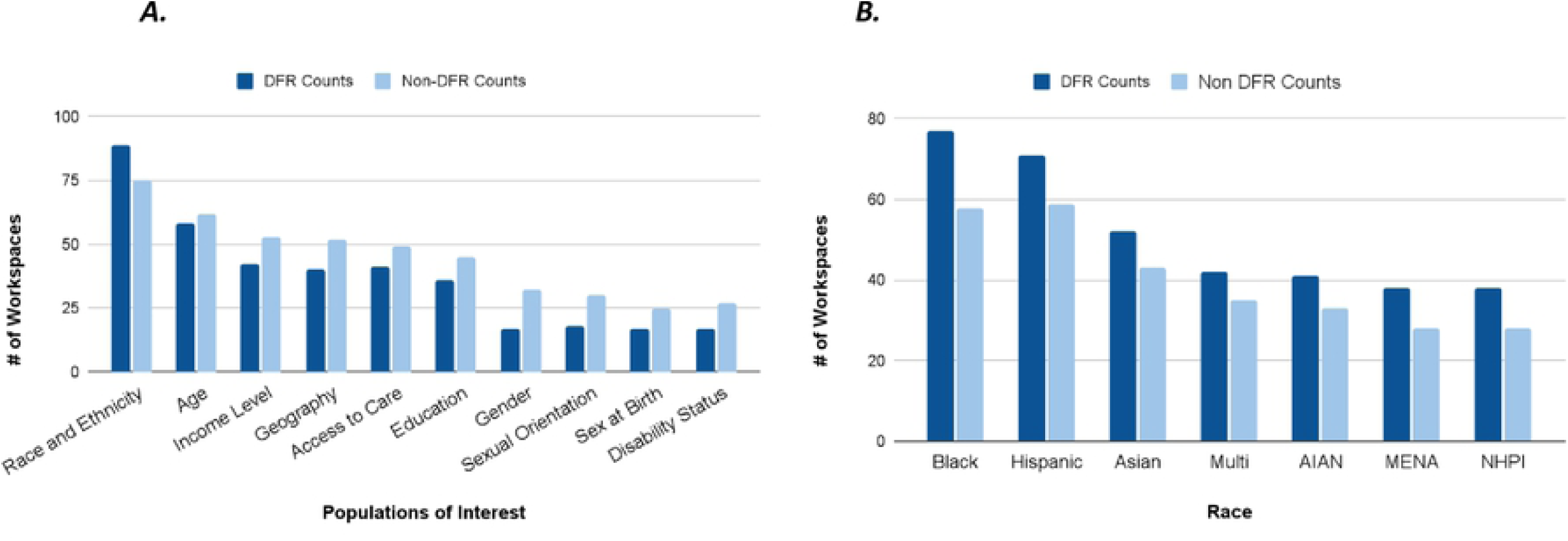
Comparing Populations of Interest. Comparing DFR to non-DFR in Panel A, only race and ethnicity was statistically significant (Panel A, p = 0.019). The distribution of the race and ethnicity categories in DFR compared to non-DFR workspaces was not statistically significant (Panel B). Black/African American, Hispanic/Latinx/Spanish, and Asian American populations were the most commonly studied across workspaces.

## DISCUSSION

Population and public health is the most common research purpose across workspace types. However, genetic and social behavioral workspaces are more common in DFR, whereas educational and methods development or validation workspaces are more common in non-DFR. Interestingly, the top 5 disease categories studied were cardiovascular disease, brain and mental health disorders, cancer, reproductive disease, and immunology, even when subsetting to workspaces doing genetic research. This suggests that a majority of researchers in this initial cohort (Dataset C) are interested in studying these major disease categories, regardless of whether they intend to leverage genomic data in their analyses. Focusing on cardiovascular disease and cancer workspaces, these workspace descriptions could be further classified based on particular Athena OHDSI catalog terms. One of these subcategories was “not primary focus” indicating that while a term was present somewhere in the workspace description, this disease focus was not the primary goal of a workspace research project. As shown in the supplemental figures, the non-primary focus subcategories in cardiovascular disease and cancer workspaces were significant in counts. Besides this finding, for the most part, the manual selection of disease categories is successful using the Athena OHDSI catalog (Figure S2, Figure S3).

Additionally, focusing on current, active workspaces limits the ability to perform retrospective analysis of a larger scale by excluding some of the original demonstration and practice workspaces. Another limitation or possible source of bias is the manual categorization of which conditions are included in the disease categories. We attempted to correct for some of this bias through use of the Athena OHDSI catalog for the disease category selection. Using R is a more systematic, replicable approach to the analysis as well, ensuring that the methods used were able to be done on each of the datasets. Emphasizing researcher transparency and retrieving early workspace data may help mitigate some of these limitations. However, the scale is yet limited by the novelty of the Researcher Workbench as a tool, which opened for beta testing in May 2020, and by the compact timeframe for data inclusion of April -June 2021.

Future work will be able to advance these findings with more systematic approaches, and with time, the ability to perform more comprehensive, longitudinal analyses. Additionally, later analyses of the workspace descriptions can continue to standardize the curation of disease categories by using natural language processing or artificial intelligence methods to eliminate some of the inaccuracies or human biases with creating these categories. Study of changes in the workspace descriptions, and thus, the Researcher Workbench over time, can also be performed. Overall, this research highlights analysis of an initial cohort of the Researcher Workbench workspaces, identifies major disease categories, research purposes, and populations of interest, and can help us continue to grow and shape the *AoU*RP platforms in a way that benefits both participants and researchers interested in that patient data. Together, future research of this type will continue to support the *AoU*RP goals of creating a representative participant cohort as well as facilitate access of this data by a diverse group of researchers.

## Data Availability

All .R and .csv files for the figures generated from the data are available on the GitHub repository: https://github.com/rkerawala/AoURP-RWWs

## ACKNOWLEDGEMENTS

We would like to acknowledge Brandy M. Mapes at Vanderbilt Medical University Center for her assistance in ensuring this manuscript met AoURP standard guidelines. We would like to acknowledge Francis Ratsimbazafy and Michael Holmes for their contributions to data analysis. We would also like to acknowledge Romina Foster-Bonds and Jill Waalen for their feedback and review of this manuscript. The All of Us Research Program would not be possible without the partnership of its participants.

***S1. Disease Categories—Genetic Workspaces***. *Over time, the top 5 disease categories were cardiovascular disease and related ailments, followed by brain and mental health disorders, cancer and tumors, reproduction, and immunology. Overall, the same categories were studied in genetic workspaces compared to all of the workspaces. None were statistically significant over time (Fisher’s Exact test, p > 0*.*05)*.

***S2. Disease Categories—Cancer Subtypes Distribution***. *Notably, “cancer” was included in a workspace, as a non-primary focus most often within the cancer category. This was followed by miscellaneous cancer types, cancer overall, and cancer in combination with other diseases, that varied by each individual workspace*.

***S3. Disease Categories—Cardiovascular Disease and Related Disorders Distribution***. *Notably, cardiovascular disease in conjunction with other diseases was a primary focus in the majority of workspaces from this category. Other major subcategories included CVD, other cardiovascular-related disorders, and non-primary focus on cardiovascular diseases*.

